# The effect of hypersensitivity pneumonitis guideline on the pathologic diagnosis of interstitial pneumonia

**DOI:** 10.1101/2021.05.30.21257203

**Authors:** Mutsumi Ozasa, Andrey Bychkov, Yoshiaki Zaizen, Kazuhiro Tabata, Wataru Uegami, Yasuhiko Yamano, Kensuke Kataoka, Takeshi Johkoh, Hiroshi Mukae, Yasuhiro Kondoh, Junya Fukuoka

**Author notes:** Corresponding author: Junya Fukuoka, MD, PhD, Department of Pathology, Nagasaki University Graduate School of Biomedical Sciences, 1-12-4 Sakamoto, Nagasaki 852-8523, Japan.

## Abstract

Hypersensitivity pneumonitis (HP) and interstitial pneumonia (IP) have several overlapping characteristics, and a high diagnostic concordance rate of HP is rarely obtained. New guideline, highly influenced by pathology, were devised for its diagnosis. We recently reported that the 2020 HP guideline may result in a possible overdiagnosis of fibrotic HP (fHP) through multidisciplinary discussion. Here, we attempted to investigate the impact of the 2020 HP guideline on the pathological diagnosis of cases previously diagnosed as IP. We classified 247 cases with fibrotic IP diagnoses sourced from 2014–2019 into three categories according to the 2020 HP guideline: typical, probable, and indeterminate HP. The original pathological diagnosis and categorization based on the 2020 HP guideline were compared. The clinical data including serum data and pulmonary function tests were compared among groups. The number of cases that changed to HP from an original diagnosis other than HP based on the guideline was 56 (23%). The clinical data of these cases bore a greater resemblance to cases diagnosed as indeterminate for HP than those diagnosed as typical or probable. The ratio of typical and probable HP to the total cases was significantly lower when using a transbronchial lung cryobiopsy (TBLC). Based on the new guideline, the pathological diagnosis of HP efficiently excluded HP cases but increased the rate of HP diagnosis for cases with fibrotic IP that may not suit an HP diagnosis. Thus, a TBLC may not be useful when imparting findings for fibrotic HP diagnosis using the new criteria.

**Key messages:**

- What is the key question? > Does the 2020 HP Guideline improve pathological diagnosis?
- What is the bottom line? > By applying the new guideline, about 1/4 of the pathological diagnoses were changed from not chronic HP to probable or typical fHP, but most of the changed cases showed a clinical picture closer to not chronic HP.
- Why read on? > To understand that simple application of the pathological criteria of the HP guideline may cause a misleading diagnosis.

## INTRODUCTION

The classification of interstitial pneumonia (IP) is vitally important for predicting a prognosis and deciding on a treatment plan.[1] However, the diagnosis of IP remains challenging even for experts specializing in respiratory diseases. Studies have reported high interobserver variability in the identification of fibrotic hypersensitivity pneumonitis (fHP).[2,3] One study, based on their own diagnostic algorithm, suggested that nearly half of the current cases of HP have potentially been misdiagnosed as idiopathic IP.[4] This shows the diagnostic variables between teams. To date, there has been no consensus on the criteria for the pathological diagnosis of fHP and most pathologists diagnose fHP based on textbook definitions and literature reviews.[5–7]

The treatment and diagnosis of fHP are difficult. HP is always included in the differential diagnosis of fibrotic IP, although the standardization of the diagnosis of fibrotic HP has been long anticipated.[8,9] Thus, the formulation of specific guideline providing diagnostic criteria for differentiating between HP and IP is highly desirable. Recently, diagnostic criteria were developed in the Modified Delphi Survey by leading authorities in this field[10] and HP guideline were published in 2020.[11]

According to this guideline, the pathological diagnoses of HP are divided into the following four categories: “typical HP,” “probable HP,” “indeterminate for HP,” and alternative. The final multidisciplinary discussion diagnosis using the suggested algorithm would lead to the identification of HP and, if the pathological criteria point towards typical or probable HP, the patient will be clinically treated for HP regardless of the clinical and radiological features. This illustrates the critical value of the pathological diagnosis.

If the idiopathic pulmonary fibrosis (IPF) and HP guidelines are used simultaneously, the decision to select the more suitable of the two guidelines would be difficult in some cases. Furthermore, fHP cannot be easily distinguished histopathologically from diseases that also exhibit patterns of usual IP (UIP) such as collagen diseases.[12,13]

Recently, we have reported on the possibility that about half of the cases diagnosed as IPF by multidisciplinary discussion (MDD) according to the fHP guideline may be overdiagnosed as fHP.[14] In the former manuscript, we also have shown that a pathological diagnosis has a significant impact on the overall diagnosis. At present, no study has investigated the influence of the histological criteria of the 2020 HP guideline on the diagnosis of IP and the extent of the correlation between the diagnostic variations and the clinical features. The present study focused on the pathological domain, applied the HP guideline to cases of fibrotic IP and compared the pathological diagnoses of the new guideline to the original pathological diagnoses to elucidate our findings.

## METHODS

The research protocol was approved by the institutional review board of our institution (No. 20101918). Patients with pathological evidence of fibrotic IP obtained using video-assisted thoracoscopic surgeries (VATS) or transbronchial lung cryobiopsy (TBLC), whose pathology was sent to Nagasaki University Hospital for consultation between 2014 and 2019 from Tosei General Hospital were enrolled in this study. Only pathological diagnoses with a synoptic report of the findings made by multiple pathologists were used, and the presence or absence of granulomas and peri-airway lesions in the described findings were identified. Patients with a clinically definitive diagnosis of collagen disease and those with a diagnosis of non-IP conditions, such as sarcoidosis and pulmonary alveolar proteinosis, were excluded.

All pathological findings from each case were reviewed and confirmed by two pulmonary pathologists (M.O., J.F.) who have dedicated expertise in interstitial lung disease. The pathological findings that were confirmed were airway-centered fibrosis, loose granulomas indicating fHP, lymphoid follicles with germinal centers, plasmacytosis, aspirated particles, and necrotizing granulomas indicating alternative diagnosis bases on the new HP guideline. Cases with those extracted findings were considered as alternative diseases and were excluded from further analysis based on the HP guideline. Our original criteria of HP diagnosis were based on the major references in the field [5–7] including more than two of the following findings: poorly formed loose granulomas, interstitial giant cells with a cholesterol cleft, peribronchiolar metaplasia, airway-centered accentuation, bridging fibrosis, and diffuse cellular infiltration.

Patients were classified into three categories: typical fHP, probable fHP, and indeterminate for fHP according to the 2020 HP guideline. The original pathological diagnosis and categorization based on the 2020 HP guideline were compared. Clinical information was extracted from all cases and a comparative study was conducted between the group that did not change between fHP and HP, and the group in which the original diagnosis changed upon application of the HP guideline. The clinical information collected were age, sex, smoking history, IgG, Krebs von den Lungen-6 (KL-6), surfactant protein D (SP-D), bird antibody, respiratory function, and bronchoalveolar lavage fluid (BALF) result.

The patients were categorized into four groups. First, cases with an original diagnosis of HP, which was further judged to be typical or probable HP according to the new guideline included in the HP+/HP+ group; second, cases whose original diagnosis was not HP and changed to either typical or probable HP according to the HP guideline included in the HP-/HP+ group; third, cases which were not originally diagnosed as HP, which were judged to be indeterminate for HP by the new guideline, which were designated as the HP-/indeterminate group; and finally, cases diagnosed as HP, which were judged to be indeterminate for fHP by the new guideline evaluation, which were designated as the HP+/indeterminate group. The effects of the HP guideline on each modality, i.e., VATS, biopsy, and TBLC, were compared. Statistical analyses were conducted using a Fisher’s exact test followed by a chi-squared test, one-way analysis of variance, and a multivariate Tukey analysis. To visualize the change in diagnosis, an alluvial diagram was created using GGalluvial in R. Statistical analyses were performed using the open-access EZR software.[15]

## RESULTS

A total of 371 out of 780 consultation cases with fibrotic IP were selected from a single hospital. We excluded 124 of the 371 cases owing to cellular IP without fibrotic change, connective tissue disease-related IP, lymphangioleiomyomatosis, pulmonary alveolar proteinosis, sarcoidosis, carcinoma, and other disqualifying findings. After the appropriate trimming of the dataset, 247 cases were enrolled in the study.

### Disease distribution based on the 2020 HP guideline

Forty-two cases were histologically classified as alternative diseases by the guideline and were excluded due to the presence of strong histological autoimmune features such as plasma cell infiltration, extensive lymphoid follicles, or sarcoid-like reaction. These pathological diagnoses included 36 IPs with autoimmune features, three unclassifiable idiopathic IPs, two sarcoid-like reactions, and one human adjuvant disease. Eventually, 205 cases were classified into typical fHP (*n* = 26), probable fHP (*n* = 59), and indeterminate for fHP (*n* = 120) using the 2020 HP guideline (Figure 1).

**Figure 1.**
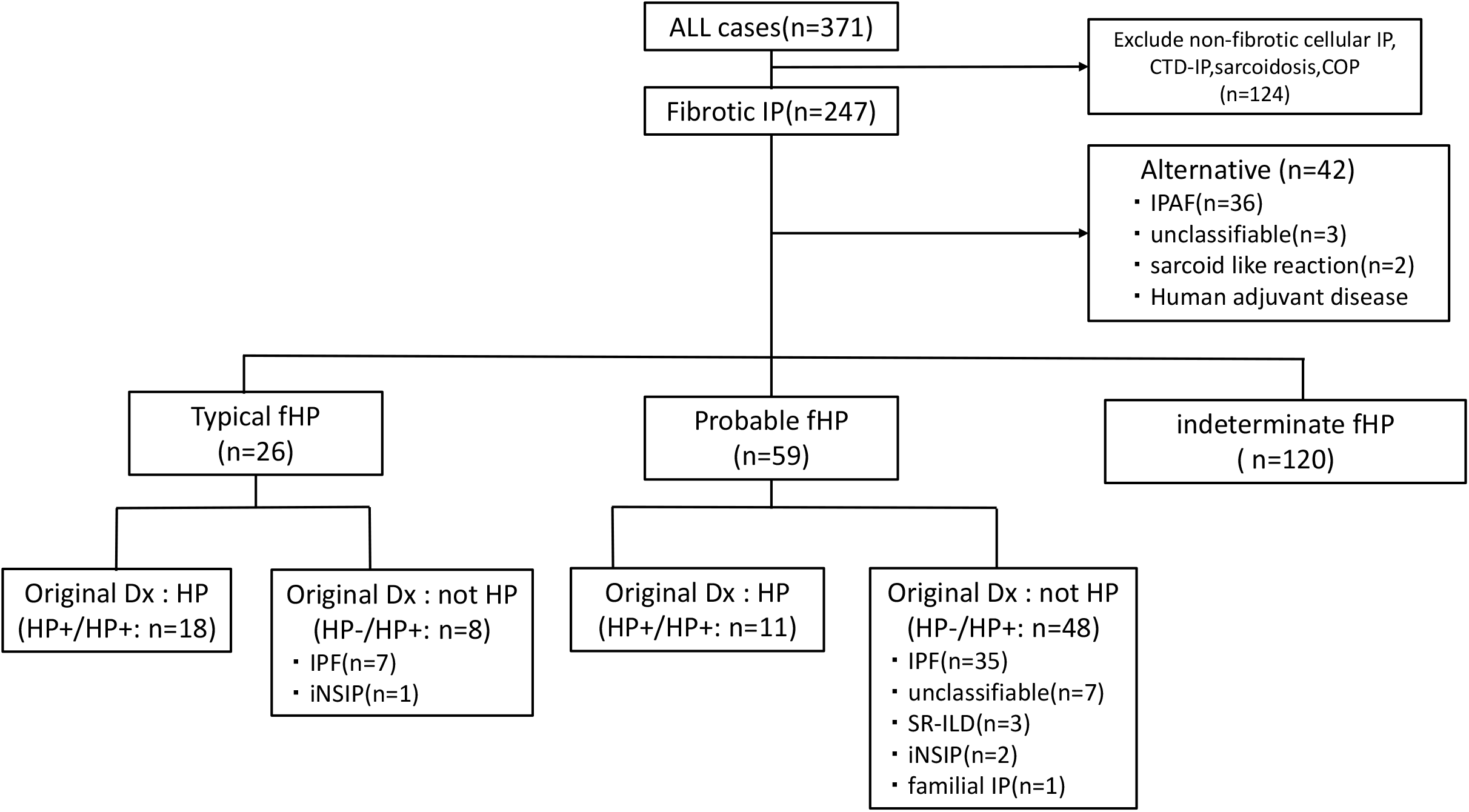
Flow diagram of the selection and application processes of the hypersensitivity pneumonitis guideline for cases with fibrotic interstitial pneumonia diagnosed using surgical lung biopsy and transbronchial lung cryobiopsy. fHP: fibrotic hypersensitivity pneumonitis, CTD-IP: connective tissue disease-associated interstitial lung disease, COP: cryptogenic organizing pneumonia, iNSIP: idiopathic nonspecific interstitial pneumonia IPF: idiopathic pulmonary fibrosis, SR-ILD, smoking-related interstitial lung disease, IP: interstitial pneumonia, IPAF: Interstitial pneumonia with autoimmune features, Dx: Diagnosis

Eighteen (70%) of the typical fHP cases were originally diagnosed with HP. Eight cases (30%) had an original diagnosis other than HP, of which seven were IPF, and one case was idiopathic non-specific IP (iNSIP).

Eleven cases (19%) of the probable fHP were originally diagnosed as HP. Forty-eight cases (81%) had an original diagnosis other than HP, among which the largest number of cases (*n* = 35) was diagnosed as IPF. The second highest number of cases (*n* = 7) was diagnosed as unclassifiable idiopathic IP; of these, three cases were diagnosed as idiopathic airway-centered idiopathic fibrosis (ACIF). Moreover, the diagnoses of smoking-related interstitial lung disease, idiopathic nonspecific IP (iNSIP), and familial IP were identified for 3, 2, and 1 case, respectively. There was uncertainty between the diagnosis of chronic hypersensitivity pneumonitis and IPF in the 12 cases of the 120 judged to have indeterminate fHP, ten of which were sampled by TBLC.

### Change of diagnosis by the 2020 HP guideline

There were 29 cases in the HP+/HP+ group. As shown in Figure 1, the breakdown of the 29 cases included 18 typical fHP and 11 probable fHP. The HP-/HP+ group included 56 cases, the breakdown of which included 8 typical fHP and 48 probable fHP. The remaining 120 cases belonged to the indeterminate group. There were zero and three cases in the HP+/indeterminate group before and after the erratum change of the guideline, respectively (Figure 2).[16] The latter three cases, including two VATS and one TBLC, were moved from the typical fHP to indeterminate fHP by the change reported by an erratum in the guideline. All three cases had non-necrotizing granulomas on the biopsy but lacked ACIF and were recategorized as indeterminate fHP (Figure 3).

**Figure 2.**
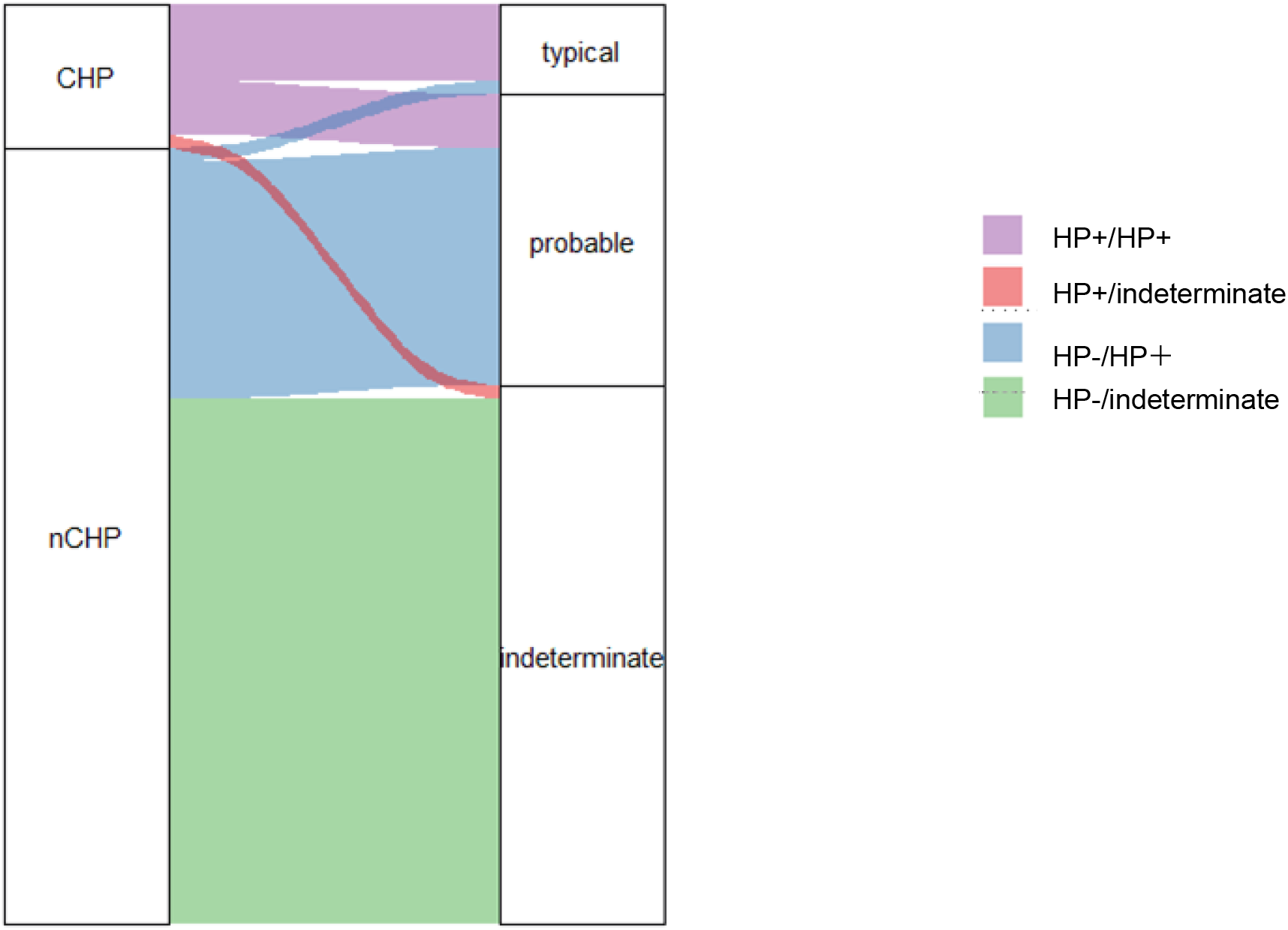
Alluvial plot to highlight the effect of the hypersensitivity pneumonitis guideline. The left and right columns show the original diagnosis and the diagnosis based on the 2020 HP guideline, respectively. Nearly one-fourth of the total cases that were originally not diagnosed as chronic hypersensitivity pneumonitis using pathology had changed to either typical or probable fibrotic hypersensitivity pneumonitis using the guideline (red colored). CHP: chronic hypersensitivity pneumonitis, nCHP: not chronic hypersensitivity pneumonitis

**Figure 3.**
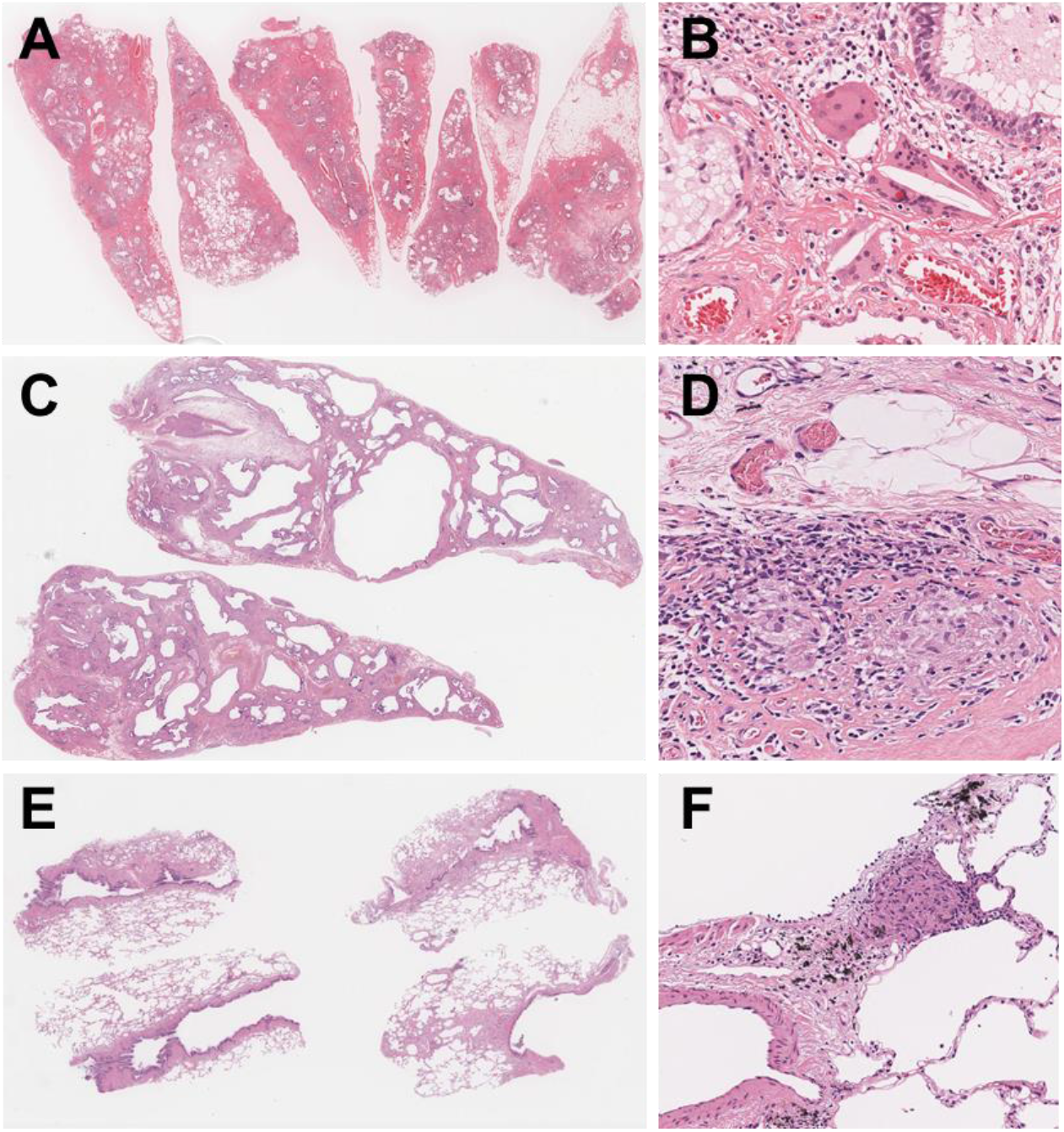
Cases recategorized as indeterminate for fibrotic hypersensitivity pneumonitis after the change of guideline by the erratum statements. A: Case of VATS biopsy shows diffuse end stage lung (H&E, x0.5). B: Higher magnification of the same case presenting an accumulation of giant cells with aggregation of histiocytes and cholesterol clefts (H&E, x20). C: Scanning view of VATS biopsy showing end stage honeycomb lung (H&E, x0.5). D: Higher magnification of the case C presenting poorly formed granulomas (H&E, x20). E: Scanning view of the case of cryobiopsy showing interstitial fibrosis. Fibrosis is found around terminal airway which represents the peripheral area inside the lung lobule (H&E, x2). F: Higher magnification of the case E showing a focus of non-necrotizing granuloma (H&E, x40).

Of the HP-/HP+ group, eight cases were converted to typical fHP. The major differential diagnosis of these eight cases was IPF (see figure, supplemental digital content 1); two of the eight cases had granulomas only in the pleura, and the changes in the lung fields were determined to be definite UIP in one case and definite fibrotic NSIP in the other. The granulomas were considered as incidental findings owing to their location. One case showed a definite UIP without conspicuous inflammatory cell infiltration. The other five cases did not have granulomas with epithelial cells, but all of them had collections of giant cells with a cholesterol cleft in the air space or interstitium. The new guideline defines multinucleated giant cells aggregate as a loose granuloma, and the location is not limited to the interstitium but can be in the air space or pleura, so it was determined to be a typical fHP. Of the 48 cases classified as probable HP by the guideline among the 56 HP-/HP+ cases, 25 possessed smoking-related emphysema (Figure 4).

**Figure 4.**
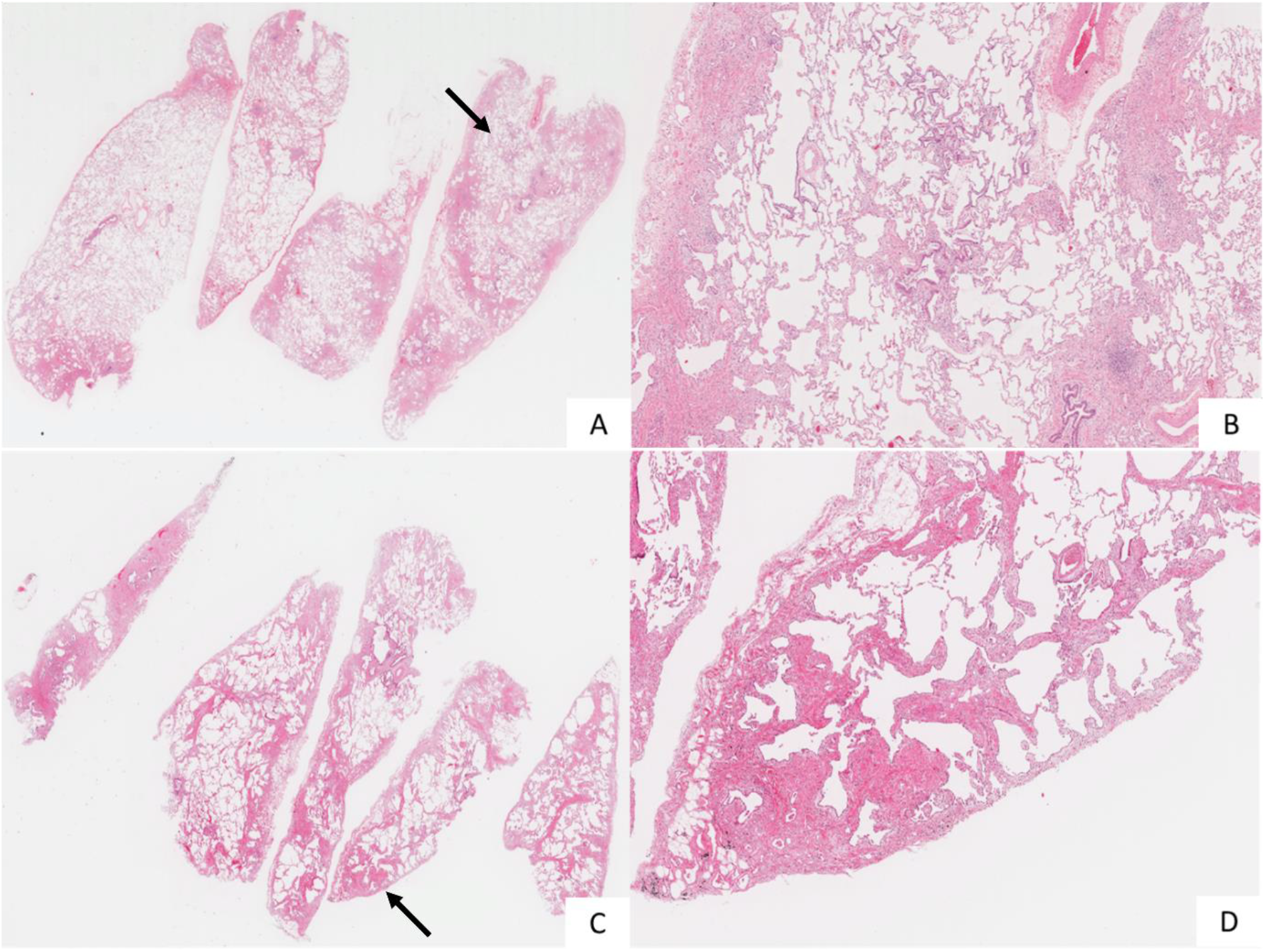
The diagnosis had changed to fibrotic hypersensitivity pneumonitis in two cases of fibrotic interstitial pneumonia using guideline. A and B. Case 1, low and middle power views (Hematoxylin and Eosin staining, 0.5× and 2×). C and D. Case 2, low and middle power views (Hematoxylin and Eosin staining, 0.5× and 4×). Both cases show patchy fibrosis accentuated to the peripheral area inside the lobule. Note the presence of airway-centered fibrosis (arrows). These cases, originally diagnosed pathologically as usual interstitial pneumonia and idiopathic pulmonary fibrosis by multidisciplinary diagnosis, fit the criteria of probable fibrotic hypersensitivity pneumonitis using the 2020 HP guideline. Case 2 shows background emphysema related to smoking.

### Correlation to clinical data

Table 1 shows the comparison of clinical information of the three groups. Factors such as KL-6 (*p* = 0.04), bird antigen (*p* < 0.01), lymphocyte fraction of BALF (*p* < 0.01), CD4/CD8 (*p* = 0.01), and % forced vital capacity (FVC) (*p* < 0.01) showed statistically significant differences.

**Table 1.** Clinical information of the three groups.

| Parameter | Original diagnosis / 2020 HP guideline diagnosis |  |  | P value |
| --- | --- | --- | --- | --- |
|  | HP+ / HP+<br>(n=33) | HP- / HP+<br>(n=57) | HP- /<br>indeterminate<br>(n=116) |  |
| **Sex** |  |  |  |  |
| Female | 10 | 12 | 40 | 0.2 |
| Male | 23 | 45 | 76 |  |
| **Age** | 61.2 ± 13 | 62.4 ± 9.2 | 64.6 ± 8.9 | 0.1 |
| **BI** | 442 ± 451 | 604 ± 520 | 504 ± 635 | 0.4 |
| **Blood parameters** |  |  |  |  |
| IgG(mg/dL) | 1580 ± 451 | 1533 ± 363 | 1489 ± 364 | 0.4 |
| KL-6(U/mL) | 2006 ± 1786 | 1191 ± 880 | 1603 ± 1332 | 0.02 |
| SP-D(ng/mL) | 359 ± 138 | 226 ± 142 | 271 ± 294 | 0.05 |
| **Bird antibody** |  |  |  |  |
| Positive | 12 | 7 | 19 | <0.01 |
| Negative | 21 | 50 | 97 |  |
| **BAL cells** |  |  |  |  |
| TC (x10<sup>5</sup>) | 2.03 ± 1.14 | 2.08 ± 1.48 | 2.26 ± 2.14 | 0.8 |
| Mφ (%) | 66.2 ± 27.1 | 80.5 ± 19.2 | 85.6 ± 84.5 | 0.3 |
| Ly (%) | 24.9 ± 24.6 | 14.2 ± 18.1 | 13 ± 16 | <0.01 |
| CD4/8 | 4.37 ± 5.57 | 3.28 ± 2.5 | 2.55 ± 1.87 | 0.01 |
| **Respiratory function** |  |  |  |  |
| %FVC (%) | 86.7 ± 22.7 | 94.2 ± 18.5 | 84.2 ± 19.4 | <0.01 |
| %FEV1.0 (%) | 83.1 ± 8.06 | 81.9 ± 9.41 | 82 ± 11.1 | 0.9 |
| %DLco (%) | 66.6 ± 21.6 | 76.0 ± 20.0 | 70.7 ± 22.4 | 0.1 |

Abbreviations: Dx: diagnosis, HP: hypersensitivity pneumonitis, BI: Brinkman index, KL-6:
Krebs von den Lungen-6, SP-D: surfactant protein D, BAL: bronchoalveolar lavage, TC: total cells, Mφ: macrophages, Ly: lymphocytes, FVC: forced vital capacity, FEV: forced expiratory volume, DLco: diffusing capacity of carbon monoxide

The serum levels of KL-6 differed significantly only between the HP+/HP+ and HP-/HP+ groups (Figure 5). The exposure to bird antigens differed significantly between the HP+/HP+ and HP-/HP+ groups and the HP+/HP+ and HP-/indeterminate groups. The lymphocyte fraction in the BALF differed significantly between the HP+/HP+ and HP-/indeterminate groups. No significant difference was observed between the HP+/HP+ and HP-/HP+ groups. CD4/CD8 in the bronchoalveolar lavage fluid differed between the HP+/HP+ and HP-/indeterminate. %FVC differed from the other parameters in that it differed significantly between the HP-/HP+ and HP-/indeterminate groups. The values of %FVC were similar in the HP+/HP+ and HP-/indeterminate groups. The %FVC of the HP-/HP+ group was significantly higher than that of the other two groups.

**Figure 5:**
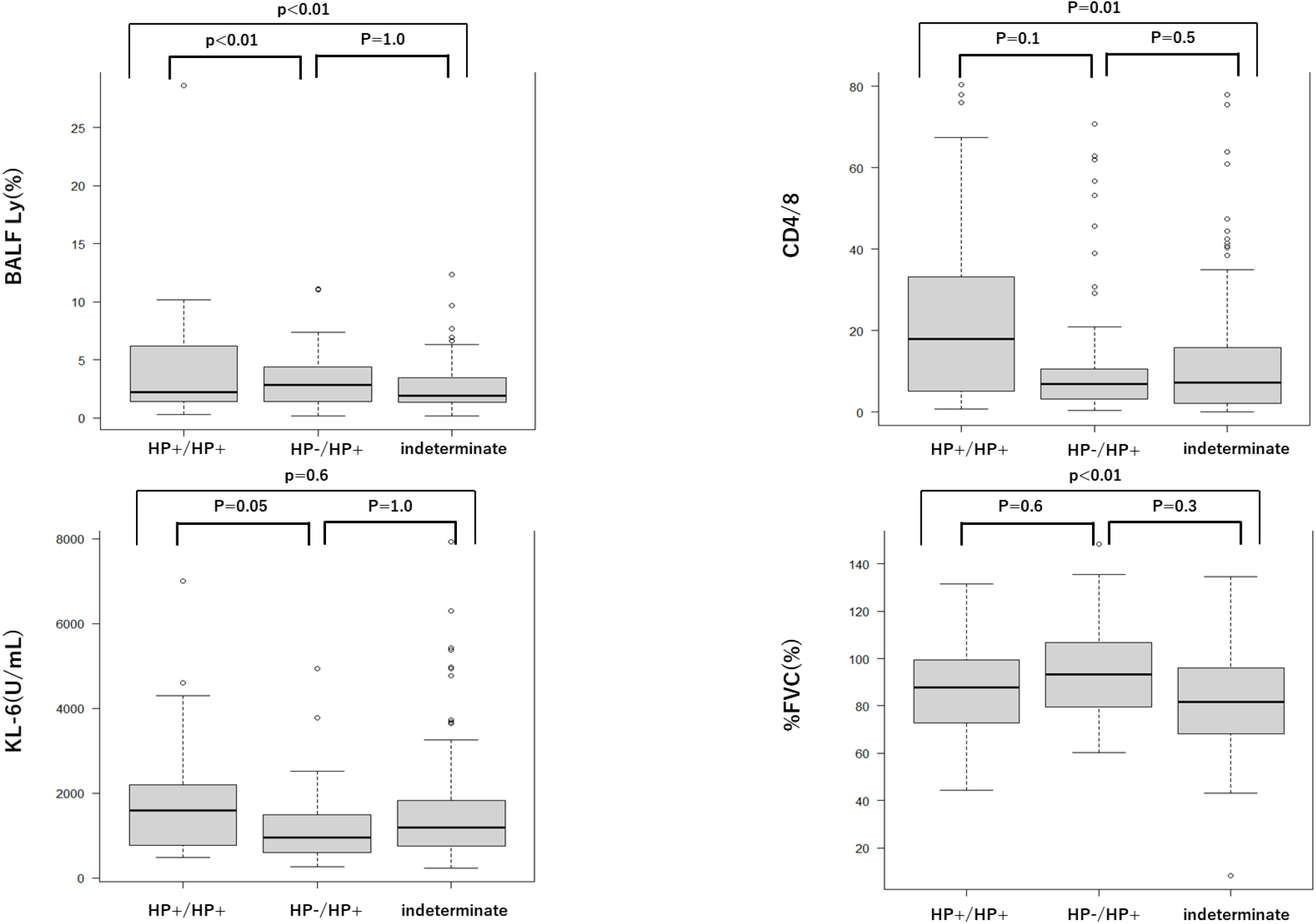
Comparison of clinical factors among three groups separated by the hypersensitivity pneumonitis guideline. BALF: Bronchoalveolar lavage fluid, Ly: lymphocytes, FVC: Forced vital capacity, KL-6: Krebs von den Lungen-6.

### Analysis by sampling modality

We also examined the differences in clinical data depending on the biopsy method used. Based on the HP guideline, 143 cases tested using VATS were classified into three categories of fHP; 23 were typical fHP, 54 were probable fHP, and 66 were indeterminate fHP. Twenty-five cases belonged to the HP+/HP+ group, 54 to the HP-/HP+ group, and 64 to the indeterminate group. A comparative study of the VATS biopsy data of the three groups revealed statistically significant differences in KL-6, bird antigen exposure, and %FVC (see table, supplemental digital content 2). KL-6 differed significantly amongst all three groups (*p* = 0.03). This finding revealed that the serum KL-6 levels were higher in fHP than those in other IPs, which was consistent with previously reported data.[17,18] Bird antigen exposure also differed significantly among the three groups (*p* < 0.01). A statistically significant difference was observed between the HP+/HP+ and HP-/HP+ groups. The %FVC also differed significantly among the three groups sampled using VATS; there was a significant difference between the HP+/HP+ and indeterminate groups (*p* = 0.02).

TBLC was performed in 62 cases, of which three were typical HP, five were probable HP, and 54 were indeterminate for fHP; there were two cases of HP-/HP+. The quality of the 62 specimens was high in confidence for 58 cases and low in confidence in only four cases, indicating that 94% of the specimens were appropriate for evaluation. The results were similar to those previously reported for TBLC specimen adequacy.[19,20] The ratios of typical HP and probable HP to the total number of VATS cases was 11.9% and 42%, respectively, and significantly lower at 4.8% and 8% for TBLC cases, respectively (*p* < 0.01).

## DISCUSSION

The 2020 the American Thoracic Society (ATS), the Japanese Respiratory Society (JRS), and the Asociación Latinoamericana de Tórax (ALAT) HP clinical practice guideline paved the way for the standardization of HP diagnosis on an international scale, which, among other benefits, is expected to prevent a misdiagnosis with other IP entities. Reporting institutional experiences on the impact of the new guideline is important to highlight its strengths and potential pitfalls. An accumulation of such reports will inevitably prompt further modifications and ensure the suitable development and evolution of the guideline. Recently, we have indicated that the 2020 HP guideline may induce an overdiagnosis of IPF cases by MDD[14]. The institutional experience presented here revealed that the 2020 HP guideline classification scheme allowed the identification of most fHP cases based on the typical and probable HP criteria. However, a pathological evaluation indicated and identified a trend of overdiagnosis of fHP by MDD. In a prior study[14], we found that pathological diagnosis holds considerable sway over the diagnosis reached through MDD. MDD is, in a sense, a method to find a compromise between different diagnostic modalities. In this process, it is possible that an erroneous conjecture from one modality may impact the final diagnosis and push it in the wrong direction. To improve diagnosis, we must seek ways to eliminate error within each modality.

In this study, 85 (34%) out of 247 cases had typical fHP or probable fHP according to the HP guideline, while fHP was not suspected in the original diagnosis, it was altered in 56 cases (22.7%) according to the 2020 HP guideline. These results suggest that the diagnosis in nearly one-fourth of all cases may be changed under the new HP guideline. Among the 85 cases, a total of 42 cases were originally diagnosed as IPF/UIP. The use of steroids treatment with or without immunosuppression in cases of IPF may be a factor that worsens the patient’s prognosis[21]. Currently, there is little evidence to determine whether those 42 patients whose diagnosis changed from IPF to fHP in the new guideline are eligible for steroids. A simple adaptation of the new guideline to these cases may lead to unfavorable outcomes.

Recently, the “CHEST guideline” for HP has been proposed by CHEST panel of experts[22]. The most important difference between the the CHEST guideline and the ATS/JRS/ALAT guideline is that the scope of alternative diagnosis that has been expanded to include IPF and indeterminate HP has been limited to only those cases that are truly unknown. This means that most of these cases will be classified as alternative diagnosis in the CHEST guideline. This content will require further study.

We compared the cases belonging to the HP+/HP+ and HP-/HP+ groups to understand the effect on cases whose diagnosis had changed to fHP using the HP guideline. We found significant differences in the lymphocyte fraction and the presence of bird antigens between them. Moreover, there was a statistically significant difference in the CD4/CD8 ratio in the BALF of the HP+/HP+ and the indeterminate groups. Although no difference between the HP+/HP+ and HP-/HP+ groups was found, the results of the HP-/HP+ group were similar to those of the indeterminate group. These data imply that the new guideline may possibly lead to a false fHP diagnosis in some cases. Further research is needed to investigate this issue.

Of the HP-/HP+ group, 48 of the 56 patients had their diagnosis changed to probable fHP owing to the presence of ACIF. This suggests that the ACIF determination is important in the diagnosis of HP. One setback may be that the HP guideline does not clearly state what level of ACIF should be considered significant. Eight other cases were converted to typical fHP, and seven of them were originally diagnosed as IPF (see figure, supplemental digital content 1). A review of their histopathology revealed granuloma or a cluster of giant cells, but they were present in the pleura and the airspace with concurrent smoking-related changes. The CHEST guideline mentions the difficulty in differentiating IPF/UIP from fHP in histopathology, and our data illustrate this well.

On the other hand, the diagnosis of IPF was made because of the high degree of fibrotic lesions on the lobes and the clear presence of honeycombing and fibroblastic focus. Proper use of both the IPF guideline and the HP guideline in such cases is challenging. In addition, strong smoking-related pathological findings such as emphysema were observed in 25 of the 56 patients in the HP-/HP+ group. It has been reported that smoking-related fibrotic lesions are found around the airways,[23–25] all of which were diagnosed to be idiopathic IP by the original diagnosis, showing that the ACIF misinterpretation should be kept in mind in cases of smoking-related pathological findings.

Before the change of the pathological criteria by an erratum, none of the cases were judged as indeterminate for fHP according to the 2020 HP guideline including cases that were diagnosed as fHP in the original diagnosis, however, three cases were recategorized as indeterminate fHP due to the absence of ACIF. As shown in the Figure 4, two VATS biopsies showed end stage honeycomb lung with granulomas, and it was impossible to determine the presence of ACIF. One TBLC was inadequate to identify ACIF, probably due to its limited size.

We examined whether the HP guideline could be applied to TBLC and found that the proportion of cases identified as indeterminate for HP was highest in the TBLC group. This is because findings such as airway-centered fibrosis and granuloma are rarely observed in TBLC without sampling a large section of lung tissue, and even if fHP is suspected pathologically, the judgment with the HP guideline is often indeterminate for fHP. This shows that the judgment of samples obtained using TBLC may be underestimated using the current HP guideline. Thus, based on our experience, simply applying the HP guideline in their current form to IP samples obtained with TBLC is not recommended.

The judgement of one criterion, ACIF, is critical for the HP guideline, however, its specificity for fHP diagnosis is unclear. Our previous study demonstrated ACIF had a high sensitivity but low specificity for the diagnosis of HP.[26] Tanizasa et al.[27] compared UIP cases with and without ACIF. Although cases with ACIF were diagnosed significantly more frequently than fHP, there was no other significant difference between them, including genetic mutations. These reports show that the clinical relevance and reproducibility of ACIF are unclear and these points need to be clarified in the future.

There are several limitations to this study. First, the study comprised a purely pathological assessment of the effect of the guideline on the HP diagnosis, and multidisciplinary discussion was not performed to reach a final clinical, radiological, and pathological consensus diagnosis. However, this was not within the scope of our study, which focused on the pathological domain, and it has been addressed in the previous study. Second, the study was retrospective in design. Third, we used cases from a single center, which may have introduced some selection bias. Nevertheless, this is the first study to date to address the impact of the newly introduced HP guideline in a large case series.

We confirmed that the pathological criteria of the 2020 HP guideline efficiently excluded most non-HP cases. Concurrently, approximately one-fourth of all cases of fibrotic IP diagnostically changed from not HP to fHP, which may not correlate with the clinical features of HP. TBLC may not impart findings for fHP, and a simple adoption of the HP guideline may not be suitable for sampling methods other than VATS.

## Data Availability

The data that support the findings of this study are available from the corresponding author, J.F, upon reasonable request.

## Acknowledgements

The authors thank Mr. Ethan Okoshi, Department of Pathology, Nagasaki University Graduate School of Biomedical Sciences, for proofreading the English manuscript.

## SUPPLEMENTAL DIGITAL CONTENT

Supplemental digital content 1. Figure.

Histopathology of cases that were not HP before the implementation of new guideline but were changed to typical fibrotic HP by these guideline. Case 1: (A) Uniform temporal pattern of fNSIP with mild destruction of alveolar structures. There is a slight PBM around the airway, this case can be concluded that there is an element of suspicion for HP (B), however there are emphysematous changes due to strong smoking in the background (C), and it is possible that the airway centered fibrosis was also caused by smoking. In D and E, multinucleated giant cells and poorly formed granuloma are seen mainly in the pleura. There was no granuloma in the lung interstitium, and the lesion was judged to be fNSIP with prominent smoking-related lesions, but the guideline judged it to be typical HP. Case 2: (A) Fibrotic lesion with structural alterations centered on the lobular margins. It is adjacent to a normal lung and shows evidence of UIP. There was a slight PBM (B) and poorly formed granulomas (D and E) in the interstitium. There was RB M_φ_ in the air space, and this case also showed smoking changes in the background. There was no strong airway inflammation or lymphocyte-based inflammatory cell infiltration, and the original diagnosis was IPF/UIP. The granuloma was judged to be non-specific, but was classified as a typical HP by the new guideline. Case 3: (A) There was fibrotic lesions with structural modifications on the lobular margins and some microscopic honeycombing with internal mucus, indicating UIP. The original diagnosis was IPF/UIP because there were no obvious airway lesions, no inflammatory cell infiltration, mainly by lymphocytes, and no poorly formed granulomas with epithelial cells. The diagnosis was changed to typical fHP by new guideline.

Supplemental digital content 2. Table. Comparison of clinical information among three groups in VATS cases.

