## Supplementary figures and images for "The effect of hypersensitivity pneumonitis guideline on the pathologic diagnosis of interstitial pneumonia"

### Supplemental Figure1

#### Slide 1
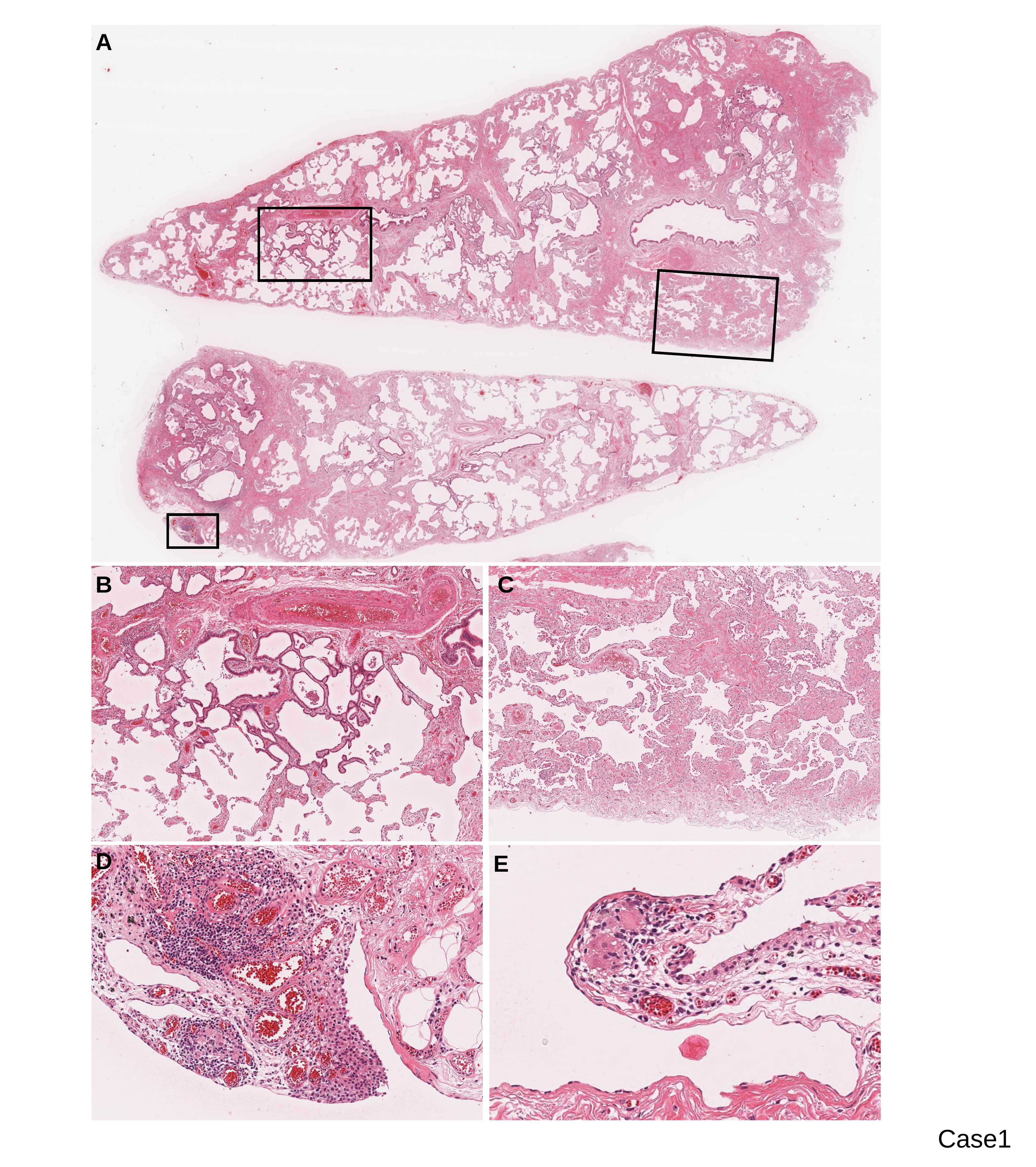

A
B
C
D
E
Case1

#### Slide 2
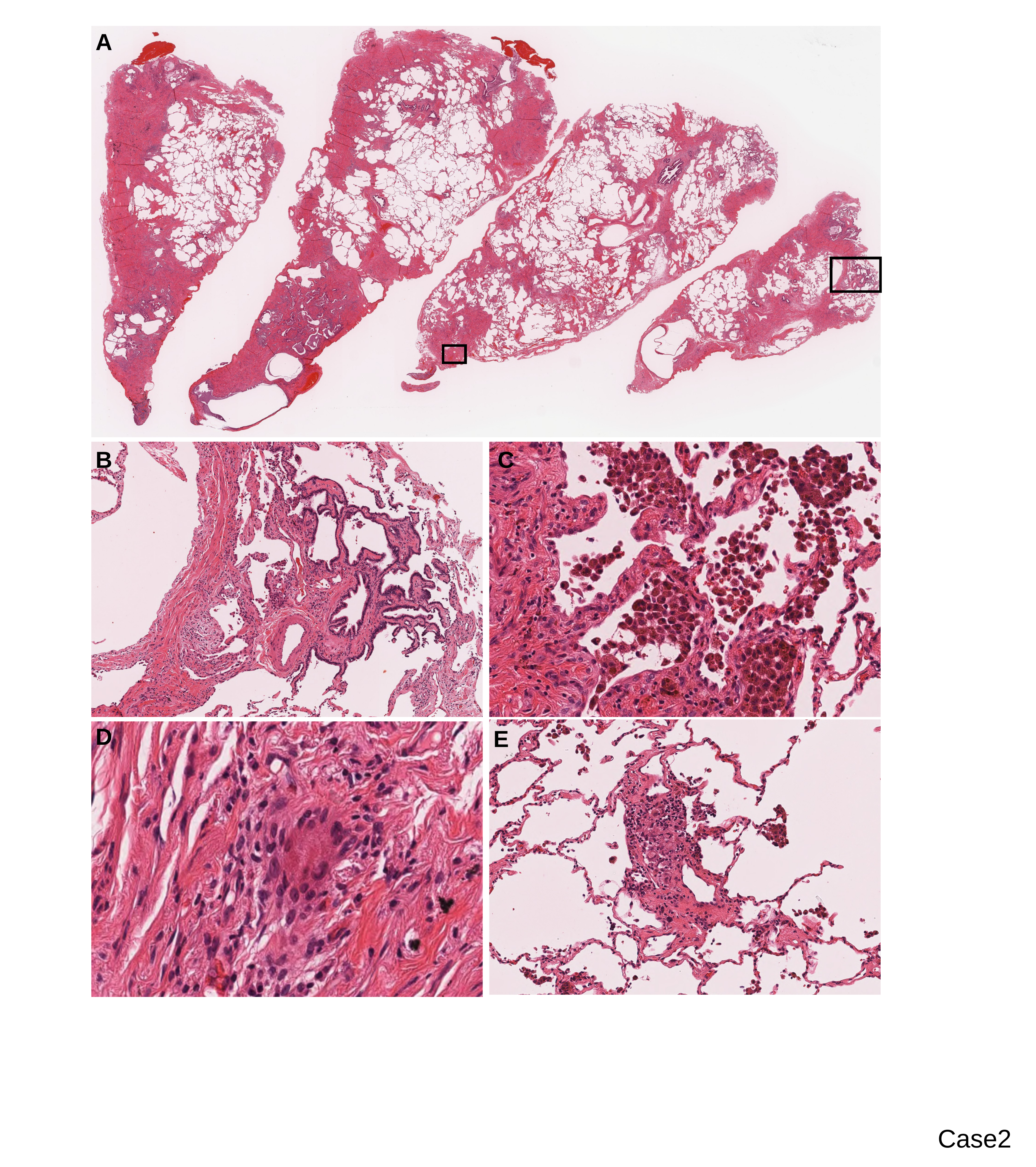

A
B
C
D
E
Case2

#### Slide 3
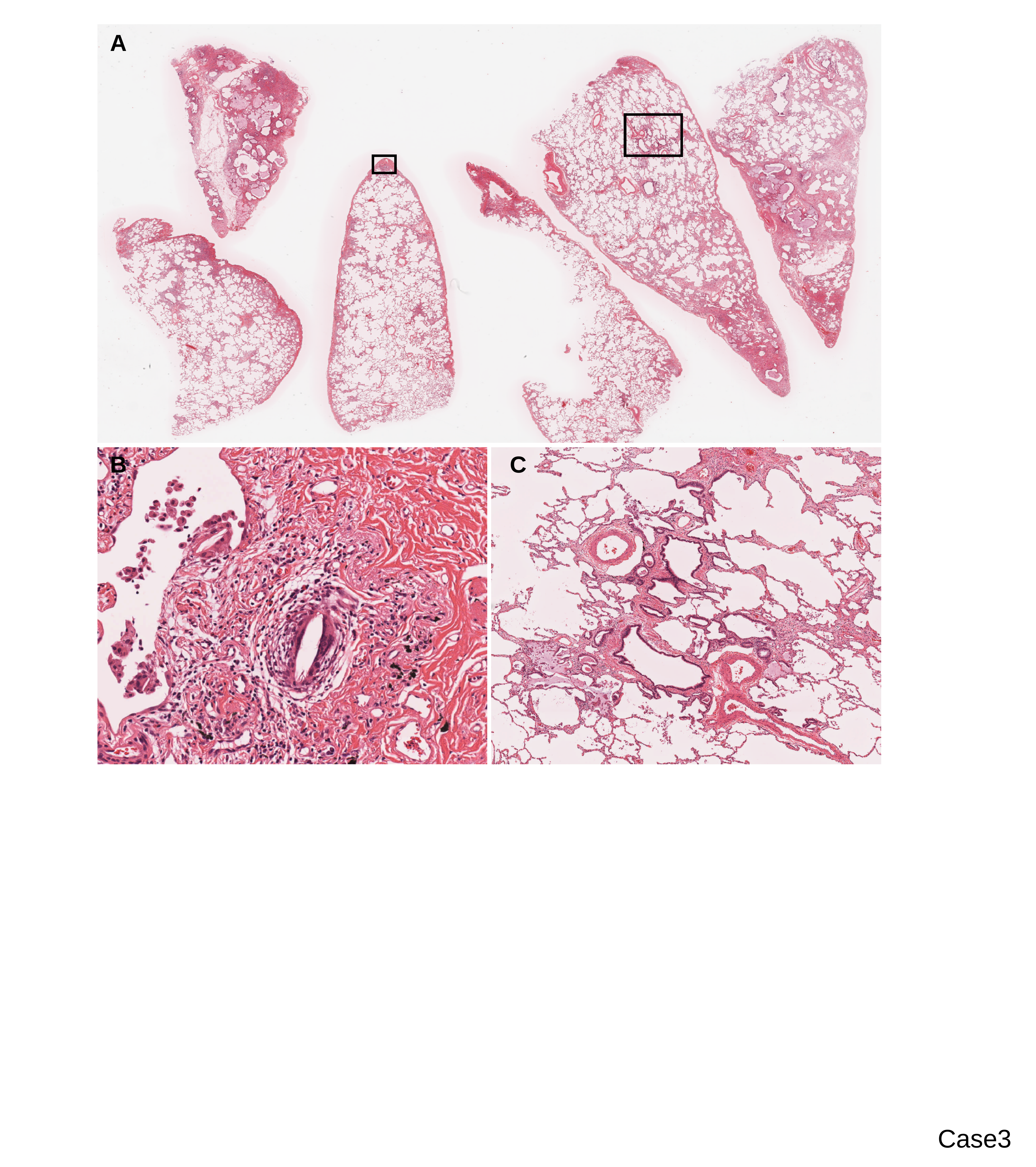

A
B
C
Case3
